# Statistical and agent-based modelling of the transmissibility of different SARS-CoV-2 variants in England and impact of different interventions

**DOI:** 10.1101/2021.12.30.21267090

**Authors:** J. Panovska-Griffiths, B. Swallow, R. Hinch, J. Cohen, K. Rosenfeld, R. M. Stuart, L. Ferretti, F. Di Lauro, C. Wymant, A. Izzo, W. Waites, R. Viner, C. Bonell, The COVID-19 Genomics UK (COG-UK) consortium, C. Fraser, D. Klein, C. C. Kerr

## Abstract

The English SARS-CoV-2 epidemic has been affected by the emergence of new viral variants such as B.1.177, Alpha and Delta, and changing restrictions. We used statistical models and calibration of an stochastic agent-based model Covasim to estimate B.1.177 to be 20% more transmissible than the wild type, Alpha to be 50-80% more transmissible than B.1.177 and Delta to be 65-90% more transmissible than Alpha. We used these estimates in Covasim (calibrated between September 01, 2020 and June 20, 2021), in June 2021, to explore whether planned relaxation of restrictions should proceed or be delayed. We found that due to the high transmissibility of Delta, resurgence in infections driven by the Delta variant would not be prevented, but would be strongly reduced by delaying the relaxation of restrictions by one month and with continued vaccination.

## 1. Introduction

Severe acute respiratory syndrome coronavirus 2 (SARS-CoV-2), the virus causing COVID-19, has continued to spread in England throughout 2020 and 2021. Spread was facilitated by the emergence of new viral variants such as B.1.177, B.1.1.7 (Alpha) and B.1.617.2 (Delta), which have dominated in late 2020 and early 2021 (Figure 1(a)-(b)). By July 2021, over 4.2 million confirmed cases and over 122 thousand deaths related to COVID-19 had been reported in England [1].

**Figure 1.**
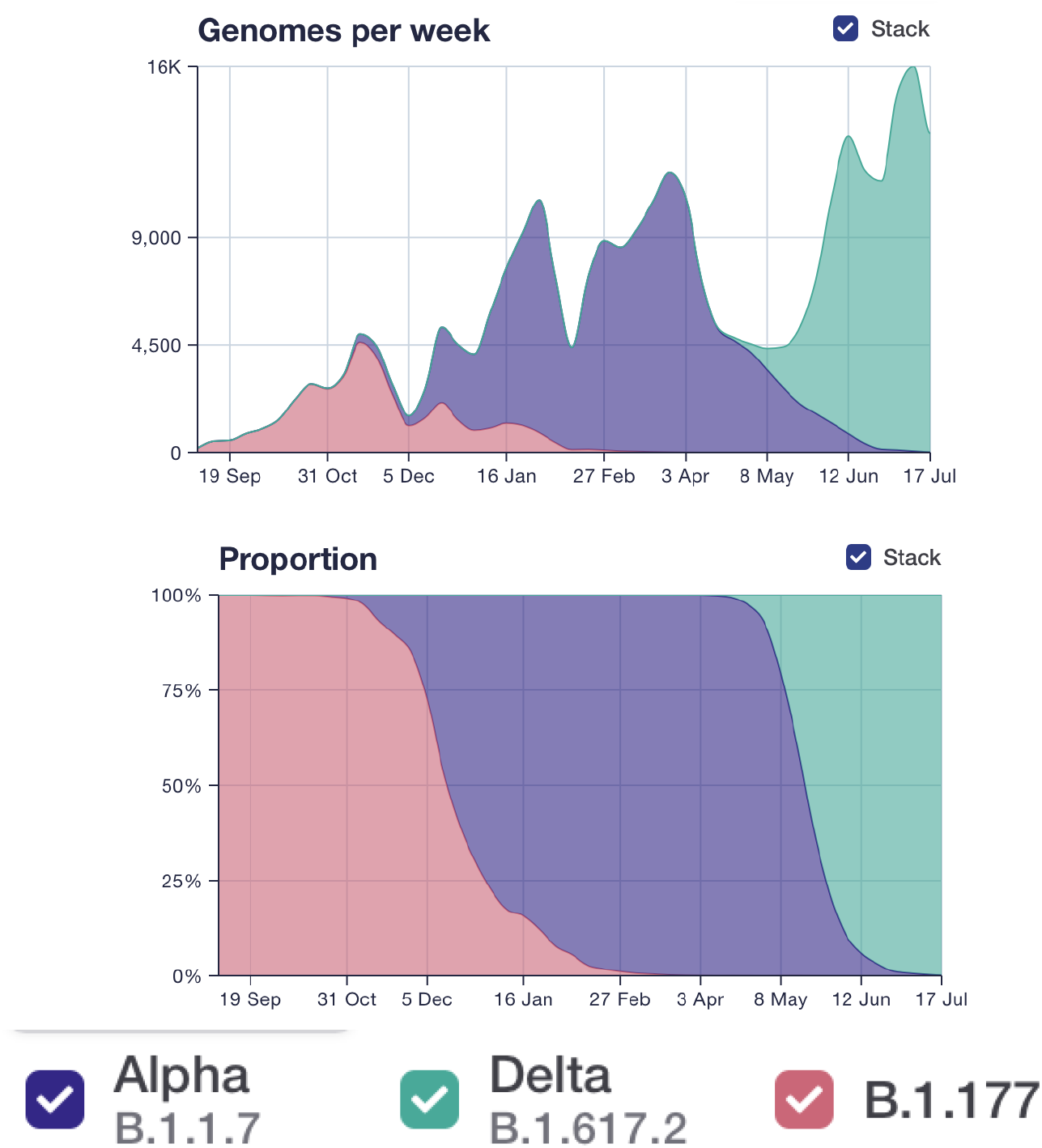
The frequency of different SARS-CoV-2 variants over the study period from COG data [3], (a) as a absolute numbers, (b) as proportions of the total.

The first wave of infections in 2020 in England was mostly of the D614G variant [2]. It was suppressed by the first national lockdown between March and June 2020 (Figure 2(a)); by June 2020 the number of infections caused by D614G were declining (Figures 1 and 2(a)). Other viral variants subsequently emerged [3], notably B.1.177 which first appeared in England in late summer 2020 and was the dominating variant over early autumn (Figure 1(a)). The Alpha variant or B.1.1.7 was first detected in South East England at the end of September 2020, and had become the dominant variant by late November 2020 (Figure 1). Unlike other strains circulating during autumn 2020, the Alpha variant had spread widely nationally by late December 2020, leading the UK Government to impose a national lockdown from January 04, 2021 [4].

**Figure 2.**
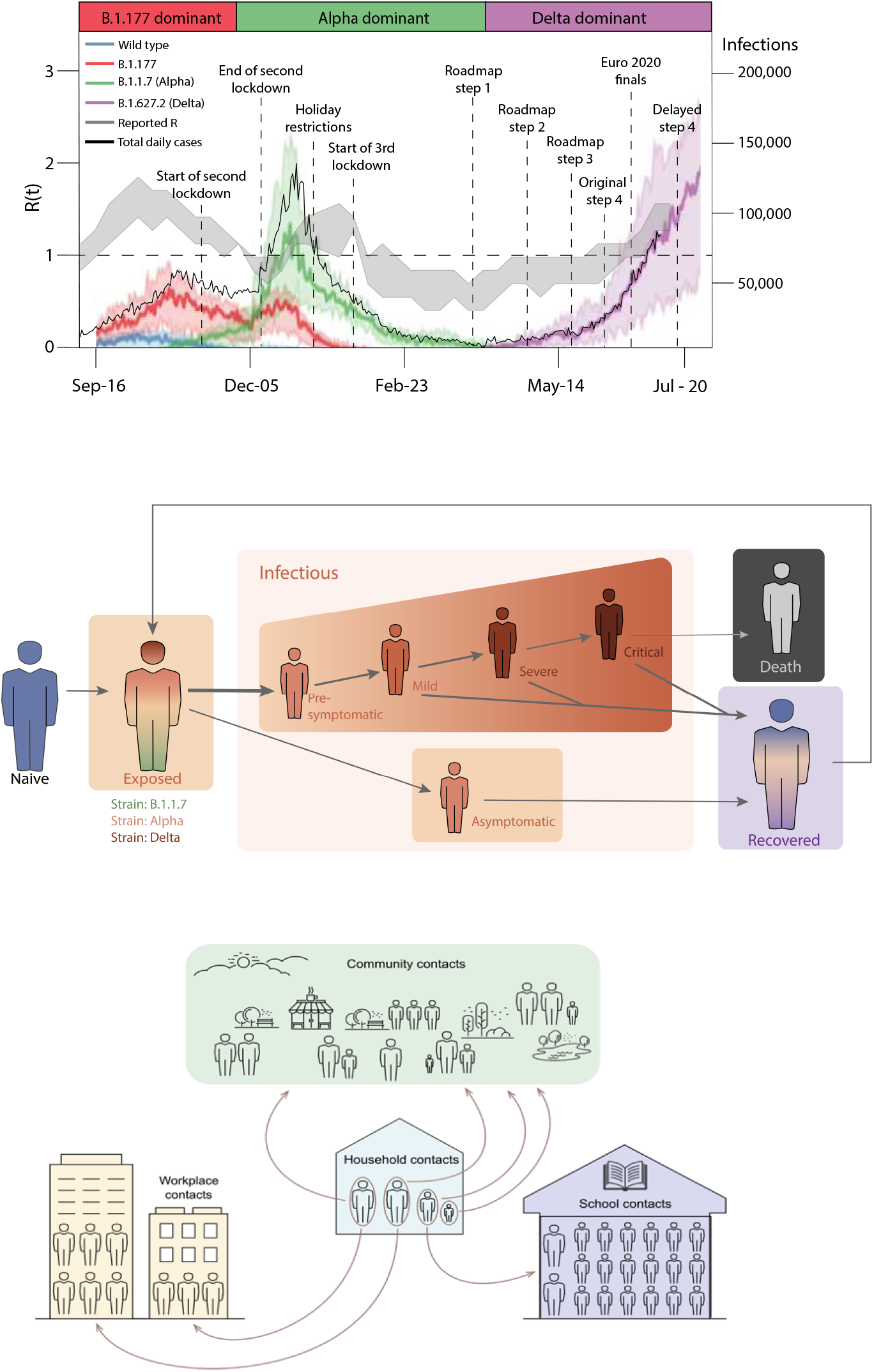
a) Model-generated daily infections by different SARS-CoV-2 variant type 9red, green and purple lines), together with data on the total number of daily cases (black line; all on the right y-axis)) and the reported R value (grey band and on the left y-axis)), in England between September 2021 and June 2021. Bold coloured lines show the median over 100 simulations, and the shaded intervals around these show the 90% confidence interval across the simulations. b) and c). Schematic of the Covasim model. (b) and of the different layers within society modelled (c). Figures (b) and (c) are reproduced with permission from [9].

In addition to this third national lockdown, an age-prioritised COVID-19 mass-vaccination programme was deployed across England from December 2020. On July 12, 2021 80% of the total population in England had received one dose and 61% of the total population had received two doses of vaccine [1].

As a result of the third national lockdown, and the successful vaccination strategy in early 2021, there was a decline in COVID-19 cases and a reduction in the effective reproduction number R below 1 over January-February 2021 (Figure 2(a)). In late February, the UK Government therefore announced a “Reopening Roadmap” for 2021 [5] schematically shown in Figure 2(a). The roadmap comprised four consecutive steps of relaxing COVID-19 restrictions alongside a continual mass-vaccination strategy as the planned exit strategy out of the COVID-19 epidemic. Step 1 on the roadmap was the reopening of schools from March 8 and relaxing of the “stay at home rule” from March 29. Steps 2 and 3 followed from April 12 and May 17 with further restrictions relaxing slowly.

Step 4 on the roadmap, with full relaxation of the social distancing measures under “back to normal” scenario, was originally planned for June 21. However, during the weeks leading up to this date, concern grew over the safety of relaxing restrictions due to rising rates of infection linked to the Delta variant. The Delta variant or B.1.617.2 was first detected in India and began spreading in England from the middle of April 2021, eventually becoming the dominant variant. There was a notable increase in COVID-19 cases and hospitalisations with COVID-19 ahead of the planned date for Step 4: between May 23, 2021 and June 17, 2021, daily reported cases rose from 1,734 to 9,371 and daily hospitalisations rose from 114 to 242 [1]. (The number of deaths from COVID-19 remained low: 7 on May 23, 2021 and 6 on June 17, 2021.) Previous studies have suggested that the rapid sweep of the Alpha variant from December 2021 was due to it being much more transmissible than previously circulating variants [6,7]. Similarly, the increase in cases and hospitalisations in June 2021 has been attributed to possible higher transmissibility of the Delta variant compared to previous circulating strains [8], although behavioural effects, for example linked to the Euro 2020 football tournament which started in June 2021, may have also played a role.

Here we report on analyses performed in June 2021 that were used to help decide whether Step 4 on the roadmap should proceed as planned or be postponed. We used statistical analysis of genomic surveillance data [3] and the agent-based model Covasim [9] to evaluate the growth advantage of the B.1.177, Alpha and Delta SARS-CoV-2 variants compared to previously circulating variants in the period between September 2020 and July 2021, and explored the epidemic trajectories in England in the first half of 2021. We illustrated the application of the calibrated Covasim model across different scenarios quantifying the impact of the vaccination strategy and of the Delta variant on planning cases, hospitalisations and deaths alongside the four steps of the roadmap in England in the first half of 2021.

This study extends existing Covasim work by explicitly modelling different SARS-CoV-2 variants, vaccination against COVID-19 in early 2021, and the impact these had on the epidemic in England.

Our results, alongside those of other modelling groups [10] were used to scientifically advise the UK Government to delay Step 4 till July 19.

## 2. Methods

### (a) Statistical analysis to quantify variants transmissibility

To assess the relative transmissibility of different SARS-CoV-2 variants in England we used publicly available sequencing data from the COG-UK Consortium [3] between September 2020 and July 12, 2021. Multivariate regression analyses quantified the change in growth rate in Alpha and Delta relative to the previous dominant variant B.1.177, which dominated between September 2020 to November 2020 (Figure 1). The count of each variant in each of the 311 lower tier local authorities (LTLAs) was modelled as a function of multivariate smooth functions across latitude, longitude and day.

The variation in spread across variants was tested using a hierarchical generalised additive model (HGAM) following the method in [11]. GAMs have the general regression form:

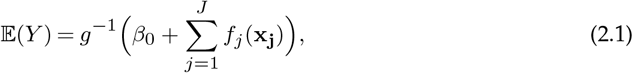

where 𝔼(*Y*) is the expected value of the response (count) assuming an appropriate distribution with link function *g*(.), *β*_0_ is an intercept, and *f*_*j*_ is a smooth function (often a spline) of covariate(s) x_**j**_.

In addition to standard fixed effects that are estimated in linear regression models, these non-linear regression models allow for smooth functions of covariates, and interactions between these, allowing for similarities between observations close in space and/or time. Within the regression, the variant can be treated both as a fixed factor (estimating the average transmission rate across the study domain) and/or as a random effect (where the smooth effects over other covariates are estimated separately for each variant). The HGAM approach follows naturally from standard generalised additive models (GAMs) [12] to allow variation in the smooth relationships across group levels which in this study are proxies for variants. In this method the smooth functions are functions of varying combinations of district latitudes, longitudes, date and variant.

The latitude and longitude of each district was taken from ONS data [14]. Districts that are closer together were assumed to be more similar than those further away, so spatial correlation was accounted for using the smooth effects. This can also assist with smoothing out data errors inherent in these observational studies, such as delays in reporting or variations in effort across local areas or time.

We treated each variant as a separate group and fitted a range of different models with a variety of different functional relationships, selecting the preferred model using the Akaike’s Information Criterion (AIC). The AIC penalises those models that fit the analysed data marginally better but are overparameterised, and therefore are expected to not fit well to other data.

The weekly count of variants in each local district was modelled using a negative-binomial distribution, which allows for potential overdispersion relative to the equal mean-variance relation assumed under a Poisson distribution. The count of each variant across each LTLA and week was the response variable, with a log link function between linear predictor and response in all model formulations. A variety of different linear predictors were fitted, including fixed effects, smooth effects and random effects. Following [11], we commenced the analysis with the simplest model assuming equivalence between variants and worked up to the most complex model assuming complete segregation between variants. HGAMs have the benefit over GLMMs, used for example in [6], of being able to account for a wider range of non-linear structures in the data, and also of allowing for the possibility of greater forms of variation in trends across and between variants.

We fitted the equivalent of the five models used in [11]. These were built hierarchically based on whether each model had a single smoother or whether they had joint ones across groups (variants), and whether these had the same degree of smoothness (often termed wiggliness). The descriptions of the models fitted can be found in Table 1.

**Table 1.** HGAM model descriptions, corresponding to the *f*_*j*_ functions in equation 2.1. All models contain the common intercept term, the same log-link function and a negative binomial distribution for the response variable.

| Model | Description |
| --- | --- |
| G | Single smooth function of latitude, longitude and week, fixed across variants |
| I | Fixed factor slope for each variant; smooth function of latitude, longitude and week, estimated separately for each variant and with different levels of smoothness |
| S | Smooth function of latitude, longitude and week, estimated separately for each variant, but assumed to have same level of smoothness |
| GI | As model I, but with additional smooth function of latitude, longitude and week shared across all variants. |
| GS | As model S, but with additional smooth function of latitude, longitude and week shared across all variants. |

Using the best-fit model, determined by the lowest AIC value, we quantified the average multiplicative advantage of the Alpha variant versus the previously dominating B.1.177 variant and of the Delta variant versus the previously dominating Alpha variant, following a similar approach to [6]. That is, we estimated the difference in exponential growth rates between competing variants *Δr*, which we transformed into a relative transmissibility value *R*(*t*) via *R*(*t*) ~exp(*ΔrT*), where T is the mean generation time which is assumed to be 5.5 days as in [6]. We note that this is an approximate transformation, assuming the generation time *T* distribution is a delta function concentrated at its mean. A more accurate transformation would be to write 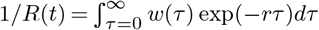, where *τ* is time since infection and *w*(*τ*) is the generation time.

To quantify this relative transmissibility value, we compared the model-fitted average trends of each variant over the time period when they started to dominate across LTLAs as a proxy for the relative progressive transmissibility. Specifically, we compared the contrast in temporal trend of the Alpha variant versus the B.1.177 variant over the period between 7th November 2020 and 30th January 2021; and of the Delta variant versus the Alpha variant between 5th May 2021 and 12th July 2021. These time periods were chosen to reflect periods when the emerging variant (Alpha and Delta respectively) were growing exponentially.

All analyses were conducted using the mgcv package [13] in R.

### (b) Dynamic modelling of the COVID-19 epidemic using Covasim

During 2020 and 2021, the stochastic agent-based model Covasim [9] has been widely used across a number of settings to track the status of the COVID-19 epidemic and to explore the impact of different non-pharmaceutical and pharmaceutical interventions. For example, in the UK, Covasim has been used to evaluate the impact of different Test-Trace-Isolate strategies when schools reopened in the UK after the first national lockdown [15], to explore the impact of wearing face coverings in schools in the second half of 2020 [17] and to simulate different scenarios of schools’ reopening at Step 1 of the roadmap [18]. Additionally, within the UK Health Security Agency Covasim is used for nowcasting epidemic metrics such as the reproduction number *R* and the growth rate *r* that track the status of the COVID-19 epidemic [19]. Internationally, Covasim has been applied in settings within the USA [20], used to inform decision making in Australia [21] and used to inform the impact of reopening borders in Vietnam [22], with a number of other studies ongoing.

In this study we extended Covasim to include transmission of three SARS-CoV-2 variants: B.1.177, Alpha and Delta in England over the period September 2020-June 2021 and also incorporated age-prioritised vaccination against COVID-19. Details of the Covasim’s core modelling framework have been published [9]; we briefly review these in section 2b(i). The specifics of modelling different variants and vaccination are contained in sections 2b(ii) and 2b(iii) with further details on modelling vaccination in [25]. We are also able to draw on social mobility data to model behaviour, with details in section 2b(iv), and can simulate different non-pharmaceutical interventions (NPIs) as described in section 2b(v).

#### (i) Covasim framework recap

In Covasim individuals susceptible to SARS-CoV-2 infection move through an exposed stage and infectious stages (asymptomatic, presymptomatic, mild, severe or critical) of infection, before recovering, getting reinfected or dying (Figure 2(b)). The model also incorporates heterogeneity in infectiousness with age. The underlying structure is “SEIRS” (susceptible-exposed-infectious-recovered-susceptible) with specific parameters describing the cross-immunity between the different variants modelled.

In this study, we used Covasim’s default parameters and the “hybrid” network structure for England (see Section 2.4 of [9] for details), with the default data included in the model for English population age structure and household sizes. The population was stratified across four population contact-network layers: schools, workplaces, households and community settings (Figure 2(c)) with pre-defined contact patterns across these layers based on the Polymod study [23].

Using Covasim version 3.0.7, we generated a population of 100,000 agents interacting over the four contact-network layers (households, workplaces, schools, and communities). Reflective of the size of the population of England, we used dynamic scaling of these 100,000 agents up to the population of around 56 million. Dynamic scaling is a standard approach within individual-based-models such as Covasim, that allows for arbitrarily large populations to be modelled whilst maintaining a constant level of precision and manageable computation time throughout [9].

#### (ii) Modelling different SARS-CoV-2 variants

In our previous work we either modelled a single strain (wild type) of SARS-CoV-2 [15,17], or implicitly modelled the effect of two strains (wild type and Alpha variant) [18]. In the latter case we simulated a single strain with time-varying infectiousness, including a logistic growth function for the relative proportion of the Alpha variant from September 1, 2020, and estimated the increased infectiousness of Alpha compared to the wild type by fitting to the increased growth in cases, in an approach similar to other work [6].

In this study, in contrast, we use new features in Covasim to mechanistically model individual SARS-CoV-2 variants by allowing different model parameters to be introduced to characterise each variant. Details of this will be explored in a separate study; briefly, we have modelled three different variants (Figure 2(a)) circulating in England in 2020 and 2021: the wild type of SARS-CoV-2 which emerged in early 2020, the B.177 variant which emerged in August 2020, the Alpha variant which emerged in late September 2020 and spread nationally between October 2020 and February 2021, and the Delta variant which emerged in late April 2021 and became the dominant variant.

The key parameters characterising these variants are a) the number of imported or seeded infections for each emerging variant at the time of emergence, b) the relative transmissibility of each variant, c) vaccine effectiveness against each variant, and d) the cross-immunity between variants.

In this analysis, the number of imported cases and transmissibility for each variant were determined during the calibration process from within the ranges derived in the statistical analysis. The assumed effectiveness of vaccines against each variant is shown in Figure 3(b) and discussed further in the next section. We explicitly modelled the ability of variants to escape immunity from prior infection or vaccination using data from binding neutralization studies [24].

**Figure 3.**
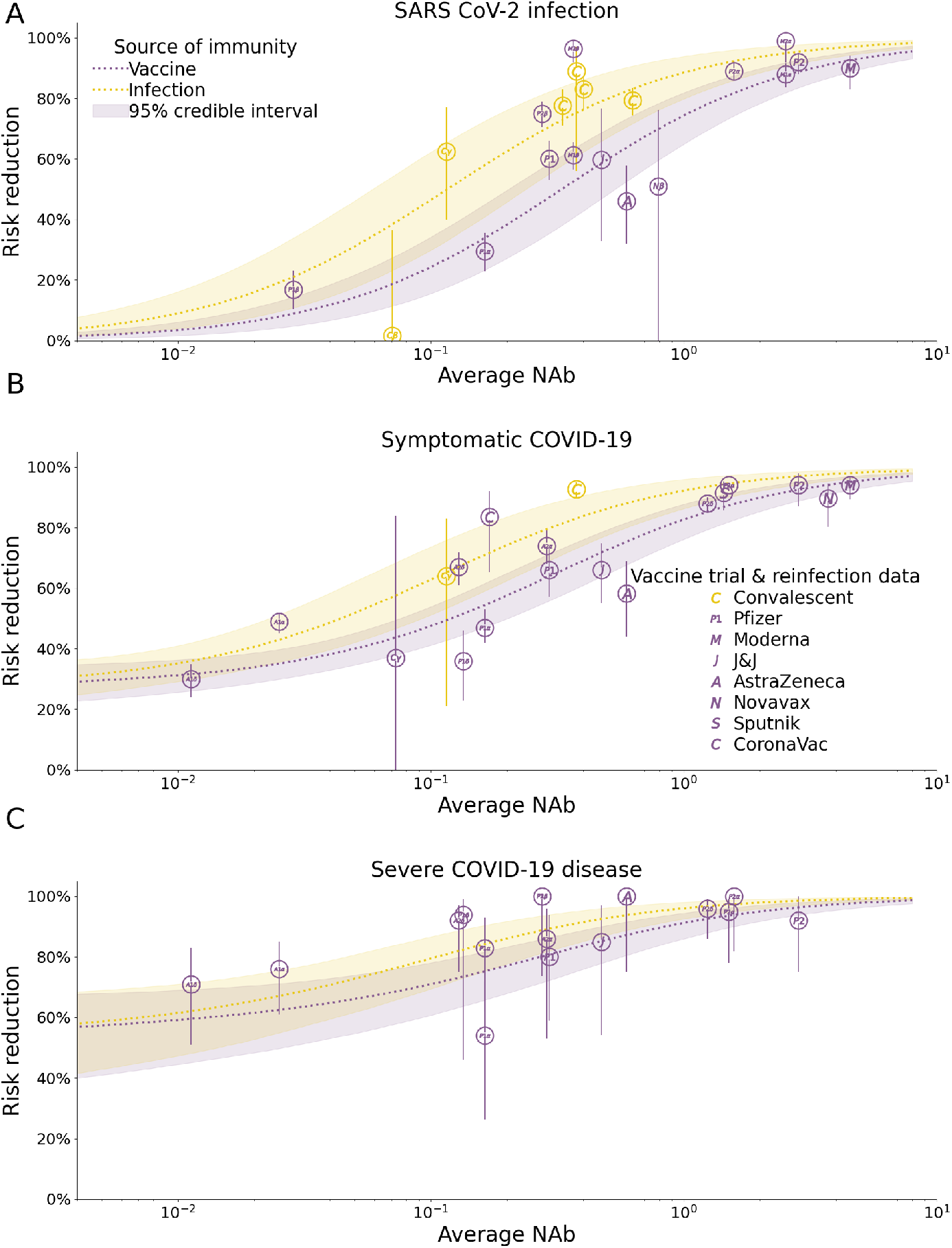
Modelled vaccine efficacy used in the Covasim model for this study. The risk reduction in infection, symptomatic COVID-19 disease and severe COVID-19 disease following vaccination or infection are modelled as functions of neutralising antibody (NAb) level. Details of the method are given in [25].

#### (iii) Modelling COVID-19 vaccination

We modelled an age-prioritised vaccination schedule with two doses given 8-12 weeks apart. Vaccines provide partial protection against SARS-CoV-2 infection, reduce disease severity, and reduce onward transmission. The schedule we used reflects the vaccination in place in England in July 2021, namely the Pfizer/BioNTech vaccine for individuals aged 65+ or under 40, and the Oxford/AstraZeneca vaccine for indivduals aged 40-64. In July 2021, the strategy was to vaccinate all adults (18+ years old) with one dose by 31 July 2021, and with a second dose within 12 weeks of the first. We did not include vaccination of those under 18 years old apart from a small proportion (10%) of this cohort with underlying conditions which would have been vaccinated under high-risk groups. Follow-up work will evaluate the impact of future inclusion of the vaccination of those under 18 years of age.

Vaccine efficacy is modelled through an immune response that primes and boosts neutralising antibodies (NAbs) in individuals and then relates the level of NAbs to protection against infection, symptomatic disease and severe disease [25–27]. The model accounts for waning NAbs over time and has been fit to vaccine efficacy and effectiveness from data in trials to date (Figure 3). For this study, we model the reported immune response induced by the Pfizer/BioNTech and Oxford/AstraZeneca vaccines as well as the ability of the Delta variant to evade vaccine- and naturally-derived immunity, which results in efficacy values within the reported ranges of these vaccines’ efficacy against Alpha and Delta [28].

#### (iv) Modelling behaviour change

Within the model we incorporated a dynamic level of transmission probability to be reflective of changes in the social mobility within different layers of society, updated weekly (Figure 2(c)). Overall levels were reflective of reported Google mobility changes [29], but scaled at different times to reflect changes such as school holidays, national lockdowns and more gradual social mobility increase during the phased reopening from March 2021 as in other studies [32]. These were necessary since the mobility changes in the Google reports stratify society in different ways to how we stratify society in the layers depicted in Figure 2(c). Next we elaborate on these.

Within the school layer we simulated children of different ages as attending either primary or secondary schools, with attendance changing due to the three COVID-19-related lockdowns and school holidays. During the third national lockdown children of key workers attended school, with estimates suggesting that around 20% of primary school students and 5% of secondary schools students were attending – an average of 14% of children [30]. For simplicity we included this in the modelling framework by simulating transmission intensity at 14% of the expected level at full attendance between January 4, 2021 and March 8, 2021. When schools reopened from March 8, 2021 we assumed a reduction in the per-contact transmission probabilities by 37% in schools i.e. simulated 63% of transmission within schools remaining from September. This was modelled as an aggregated reduction in transmission due to hygiene, mask usage and other social distancing measures in place within schools to reduce transmission, and as described in detail in [17].

For workplace social mixing, we also used Google mobility data to obtain a broad range of the change over time. Specifically, as in our previously published studies, we simulated workplace and community transmission to be 20% of their pre-COVID-19 levels during the first lockdown in 2020, and 30% during the second and third lockdown, increasing to 40% of their pre-COVID-19 levels from Step 2 i.e. April 12, 2021, to 50% of their pre-COVID-19 levels from Step 3 (May 17, 2021) and going back to 70% in workplaces and 70% in community of the pre-COVID-19 levels from Step 4 in either June or July 2021.

We modelled household transmission intensity in line with the average monthly level of increased household mobility in the Google data [29].

#### (v) Modelling non-pharmaceutical intervention

In Covasim, testing is modelled using parameters that determine the probabilities with which people with different symptoms receive a test each day, both for symptomatic and asymptomatic people, based on the reported testing levels from [1]. Tracing is modelled using parameters for the probability of reaching the contacts of those testing positive, as well as the time taken to reach them. We assume that some layers of society would be easier to trace than others; specifically, that 100% of household contacts can be traced within the same day, 80% of school and workplaces can be traced within one day, and 10% of community contacts can successfully be traced within 2 days. This results in an average of 60% of contacts traced across different layers between January and May 2021, comparable with reported monthly values from [31]. The level of adherence to isolation has parameters that differ across different layers of the population (see data sources and calibration section below). Following discussions with NHS Test & Trace, we modelled 60% efficacy of isolation over the period January 2021 and May 2021.

### (c) Data sources and calibration

For the regression analysis we used publicly available COVID-19 Genomic Surveillance data from [3].

For the epidemic trajectories, data from the UK Covid-19 Dashboard [1] were extracted consisting of daily measurements of reported cases, deaths and hospitalisations between September 1, 2020 and June 20, 2021.

We fixed the model parameters at point estimates over the period January 20, 2020 to September 1, 2021 based on values we used in our previous study [15] to fit metrics of the epidemic during this time in England. We then calibrated the model to data from the UK-COVID-19 dashboard over the period September 1, 2020 and June 20, 2021 using Optuna’s parameter sweep to find optimal values for 9 separate parameters: (a) the number of seeded infections of the 3 variants of concern (B.1.177 in September 2020, of Alpha in October 2020 and of Delta in mid-April 2021); (b) the relative transmissibility of each of the 3 variants compared to previous circulating variants; (c) the average monthly symptomatic testing rates in each of the 3 months from March to May 2021 inclusive. The calibration process was done using the Optuna (https://optuna.org) hyperparameter optimisation framework in Python to search the 9-dimensional parameter space for optimal values that minimised the absolute difference between the model’s estimates of daily cases, deaths, and severe infections (representing hospitalisations), and the corresponding data on cumulative and daily infections by date reported, deaths within 28 days, and admissions to hospital by date reported between September 1, 2020 and June 20, 2021.

This study used Covasim version 3.0.7. The code used to run the simulations across the analysis reported in this paper are available at https://github.com/Jasminapg/Covid−19−Analysis/tree/master/4_vaccines_.

## 3. Results

### (a) Results from the statistical analysis

Table 2 contains the model comparison results of the statistical analyses of the COG-UK dataset. Model GI had the lowest AIC value and hence was used for inference. All smooth terms were highly statistically significant at the p=0.05 level, calculated according to the approach of [16], implying that there are sufficient differences between the variants that models incorporating variant-specific smooth terms are required for accurate analysis. The multivariate smooth terms of latitude, longitude and day were highly statistically significant (both global and variant-specific), whereas variant-specific factor slopes were not, suggesting that simple models fitting general trends across all regions are not sufficiently flexible. This is further supported by the fact that model I had the second lowest AIC across the fitted models.

**Table 2.** *Δ*AIC values for the different HGAM formulations, that is the difference in AIC between each of the models and the best-fitting model (i.e. a value of zero corresponds to the preferred model).

| Model | ΔAIC |
| --- | --- |
| G | 10330.2 |
| I | 908.7 |
| S | 1553.2 |
| GI | 0 |
| GS | 1569.3 |

Our results suggest that the relative growth of Delta was higher than that of the previously circulating strain Alpha, hence Delta was able to outcompete other SARS-CoV-2 strains in England since May 2021 (Figure 4(a)). This is a similar pattern of behaviour to the Alpha variant, which outcompeted the previously dominating variant B.1.177 and was the dominating variant in late 2020 and early 2021, including B.1.177 (Figure 4(b)).

**Figure 4.**
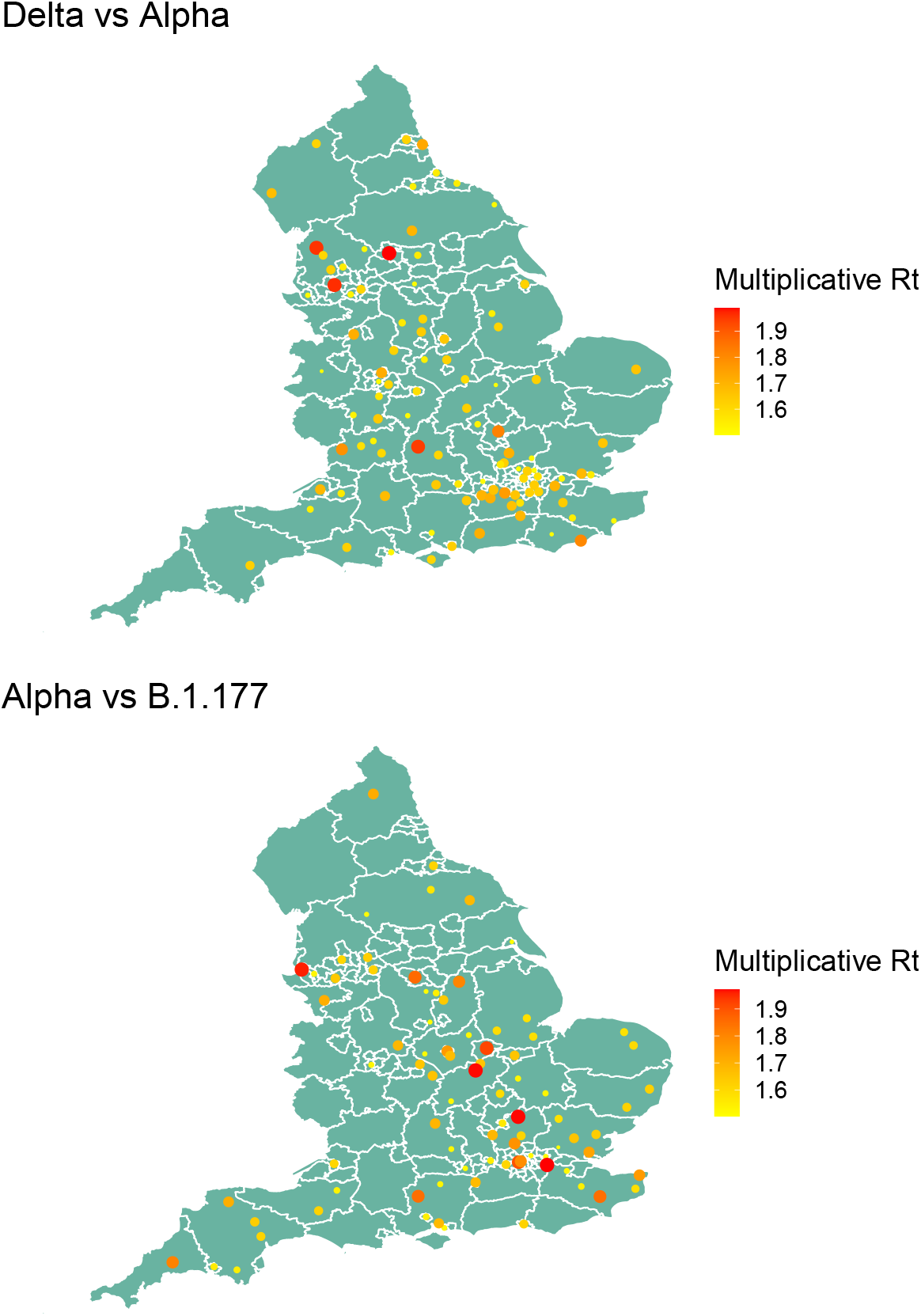
Relative transmissibility of Delta to Alpha variant (left) and Alpha variant to variant B.1.177 (right) across LTLAs for the period 5th May to 12th July 2021 (Delta vs Alpha), and 7th November 2020 to 30th January 2021 (Alpha vs B.1.177). Only LTLAs with transmissibility greater than 50% higher are shown.

The observed transmissibility of the variants across England was spatially heterogeneous (Figure 4) but Delta was consistently more transmissible than Alpha, while Alpha was consistently more transmissible than B.1.177. In LTLAs with very large (over 50%) and consistent occurrence of the Delta variant, transmissibility of Delta was 65-90% greater than Alpha, but spatially heterogeneous (Figure 4A). Similarly, the estimates for Alpha being more transmissible than B.1.177 varied between 50-80% across spatial regions (Figure 4B).

### (b) Modelling COVID-19 epidemic dynamics in England with Covasim

The model’s estimates of cases, hopsitalisations and deaths were consistent with increased transmissibility of the B.1.177, Alpha and Delta variants relative to previous circulating variants. Analogous to the statistical analysis, Delta had the highest transmissibility amongst all the circulating variants and hence has been dominating in England since May 2021 (Figure 2(a)). We found that the reported counts of cases, hospitalisations, and deaths were consistent with model estimates in which the relative transmissibilities were 1.2 ((95%CI=[1.14,1.27])) for B.1.177, 1.8 (95%CI=[1.59,1.86]) for Alpha, and 2.6 (95%CI=[2.52,2.71]) for Delta. This would imply that, on average, B.1.177 was 20% more transmissible that the dominating variants circulating in England during September 2021, Alpha was 60% more transmissible than B.1.177 and Delta was 80% more transmissible than Alpha.

Using these values for the relative transmissibility of variants, the model can effectively reproduce the epidemic trajectories of SARS-CoV-2 in England between September 1, 2020 and June 20, 2021 (Figure 5), also stratifying the number of daily infections by variant type (Figure 2(a)). Our simulations capture the increase in the epidemic as B.1.177 and Alpha variants started to spread from September 2020, with Alpha coming to dominate by the end of 2020. The third national lockdown during January and February 2021 and the vaccination programme which started in December 2020 were successful in suppressing the spread of Alpha and reduced the effective reproduction number below 1 in early 2021 (Figures 5 and 6).

**Figure 5.**
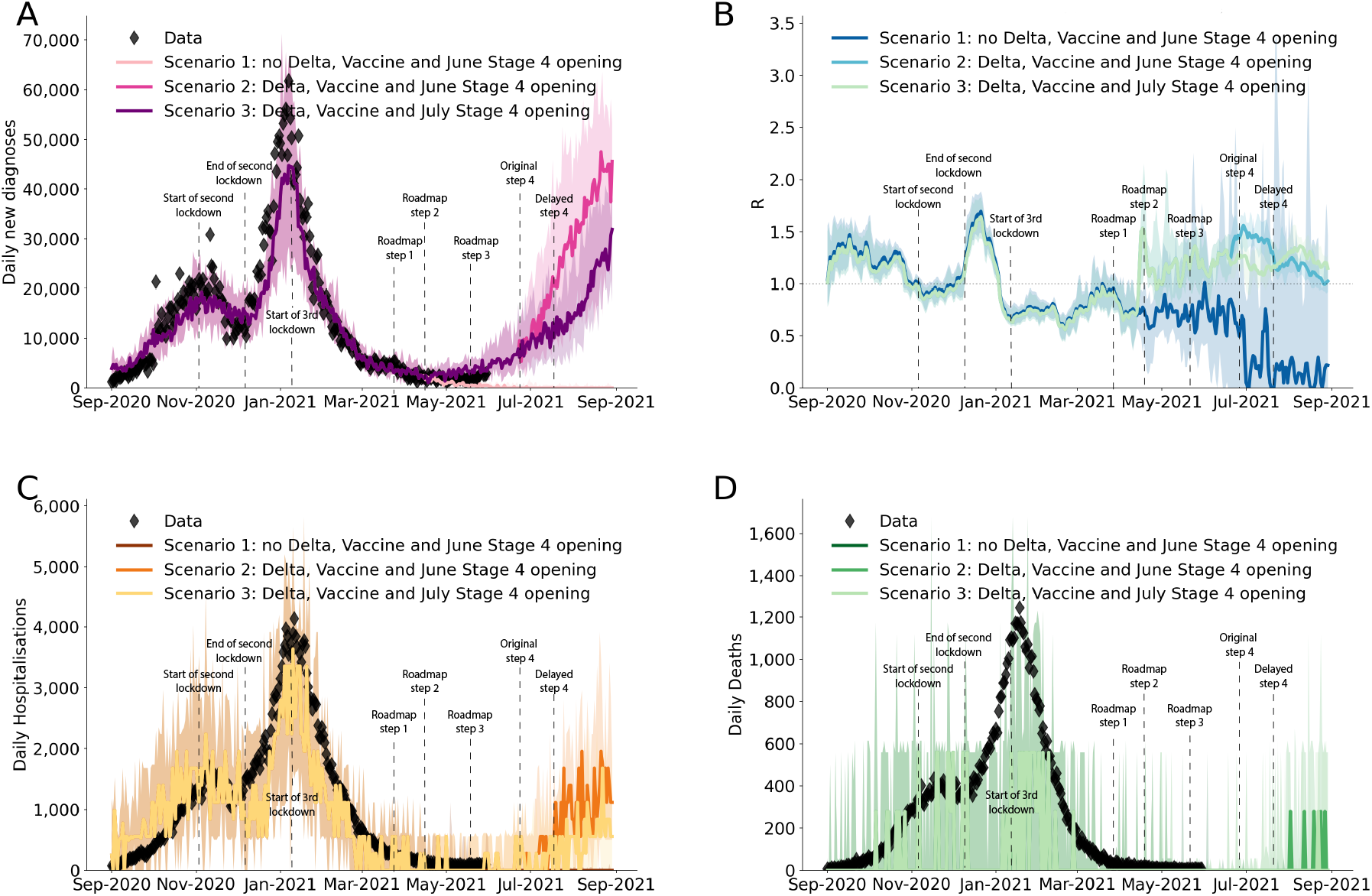
The modelled impact of delaying Step 4 of the roadmap by one month with and without the spread of the Delta variant, showing (A) the number of cases, (B) the effective reproduction number, (C) hospitalisations due to COVID-19, and (D) deaths related to COVID-19. The data from [1] is shown in diamond shapes, with the model-generated simulations overlayed. The three curves in each of the subplots illustrate the importance of Delta in continuing the epidemic spread in England after Step 3 of the roadmap, and the importance of delaying the Step 4 of the roadmap in preventing a surge in cases, hospitalisations and deaths, as well as an increase in effective reproduction number R in July 2021.

**Figure 6.**
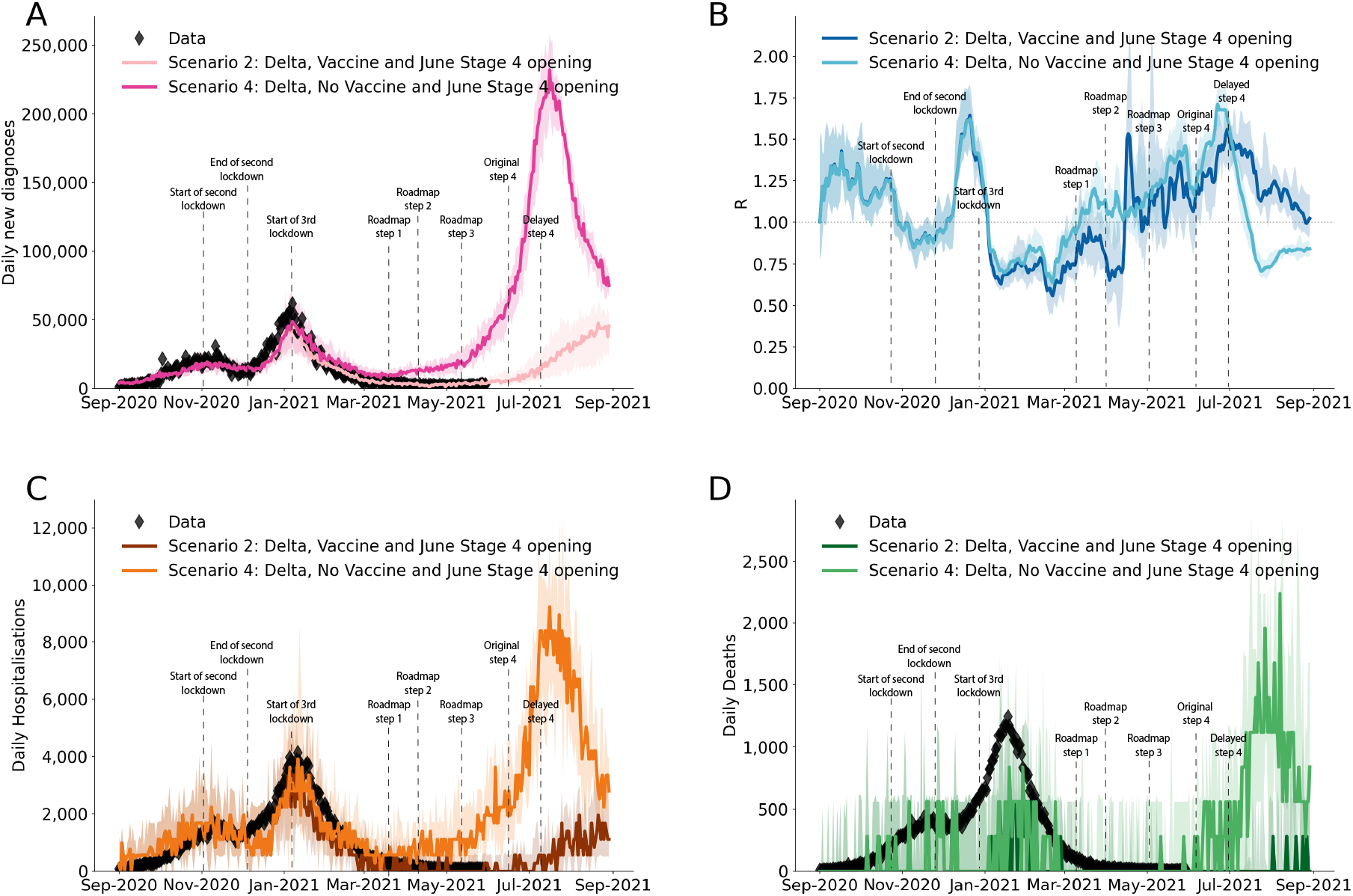
The modelled impact of vaccination against COVID-19 over the first half of 2021, showing (A) the number of cases, (B) the effective reproduction number, (C) hospitalisations due to COVID-19, and (D) deaths related to COVID-19. The data from [1] is shown in diamond shapes, with the model-generated simulations overlayed. The two curves in each of the subplots illustrate the importance of vaccination in preventing surge in cases, hospitalisations and deaths, as well as an increase in effective reproduction number R in early 2021.

Before Delta started to spread nationally, there was a period of epidemic decline with low numbers of cases, hospitalisations and deaths over the period between January 2021 and end of May 2021 (Figure 5). This allowed the phased reopening with the Steps 1,2 and 3 of the roadmap on March 8, 2021; April 12, 2021 and May 19, 2021 to occur.

### (c) Impact of delaying the roadmap by one month

After Step 3 of the roadmap, with the emergence of the highly transmissible Delta variant from mid April 2021, the number of cases, hospitalisations, and deaths started to increase despite ongoing vaccination. Our results, reported in June 2021, suggested that if Step 4 had proceeded on June 21, 2021 as originally planned, it would have resulted in a large third wave not only in cases but also in hospitalisations and deaths (Figure 5). Delaying Step 4 by one month was estimated to substantially dampen such a third wave (Figure 5).

### (d) Impact of Vaccination in presence of Delta

Although the third national lockdown played a crucial role in suppressing the spread of the Alpha variant, the roll-out of vaccination has been imperative in controlling the spread of Delta during early 2021 (Figure 6). Our results highlight a very large third wave that would have been caused by Delta had vaccination been absent in early 2021.

## 4. Conclusions

Mathematical and statistical analyses have been widely used to better understand the data and inform the science of the SARS-CoV-2 epidemic. In this paper we used these tools to simulate the spread of different SARS-CoV-2 variants in England between September 2020 and July 2021, evaluating the transmissibility of different variants relative to previously circulating ones and modelling the impact of delaying Step 4 of the roadmap in presence of Delta and with continual vaccination.

Both our statistical and our mathematical modelling confirmed that the emerging variants have been progressively more transmissible, with B.1.177 20% transmissible than previous prevailing variants, Alpha 50-80% more transmissible than B.1.177, and Delta 65-90% more transmissible than Alpha. The estimated relative advantage in transmissibility of Alpha over

B.1.177 and of Delta over Alpha from the statistical analysis was spatially heterogeneous, but the ranges agreed with the values determined by Covasim. They also agree with previous estimates, for example the transmissibility of Alpha relative to previous circulating variants was estimated as 43% to 90% by Davies et al. [6] and 50-100% by Volz et al. [7]. The transmissibility of the Delta variant versus previous prevalent variants is also close to the reported range of 69% to 83% by Sonabend et al. [8].

In agreement with other modelling results [10], our findings also confirmed that the emergence of the highly transmissible Delta variant resulted in a need to delay Step 4 of the “Reopening Roadmap” for relaxing restrictions. These results were used in June 2021 to offer scientific information of the expected consequences of delaying Step 4 till July 19. Delaying by one month was expected to significantly dampen a resurgence in infections, hospitalisations and deaths, though not prevent it completely. We also showed that while the third national lockdown in early 2021 was able to suppress the spread of Alpha, vaccination played a key role in controlling the spread of Delta, with a projected very large third wave following Step 4 of the roadmap in the absence of vaccination.

A novel feature of this work is that, whilst previous studies have shown the relative transmissibility of two simultaneously circulating variants [6–8], this study is the first study to model the sequential competition of more than two variants over different time periods. Both the statistical analysis and Covasim were able to quantify this progressive transmissibility. Understanding competition between variants is crucial for planning responses to future emerging SARS-CoV-2 variants, and the methods presented here can readily be used study this. For example in future work we intend to explore co-infections with different strains of SARS-CoV-2 and different influenza strains. Additionally, our analysis estimated the relative transmissibility of different variants using two different approaches: statistical modelling and mathematical modelling. This provided a robustness check within the same study, and generated results which are also aligned with previous studies [6–8]. Modelling has been very popular during this pandemic, and while a number of separate statistical and mathematical models have been used for inference and for nowcasting or forecasting under different scenarios, statistical and mathematical modelling have only rarely been used in parallel to untangle the same question, with some exceptions [33].

We note that Covasim is a stochastic model and as with any stochastic modelling, there is uncertainty in predicted outcomes arising from inherent stochasticity, in addition to uncertainty about the values of parameters controlling the process. The uncertainty in our predictions can be seen in Figures 5 and 6. Our results were based on taking the median of 100 simulations from a stochastic process. This uncertainty increases when predictions are made over a longer time period. Hence, in Figures 5 and 6 we only projected for 4 weeks into the future. Projecting results of any model, including ours, too far into the future based on current data is unwise due to increasing uncertainty. Importantly, estimation of future epidemic trajectories will depend on the effect of expanding vaccination to include individuals under 18 years old, and also any third dose boosters to the older population cohorts, and we will explore these in future studies.

Our analyses have illustrated the importance of delaying the Step 4 of the roadmap in England in order to reduce its’ impact on the surge of epidemic. Appropriate timing of full reduction of the COVID-19 restrictions in conjunction with the fast immunisation programme against COVID-19 have been essential in preventing large surges in the English epidemic, and this should be a lesson to other countries as they are faced with exit strategies out of lockdowns.

Our work has some limitations and aspects that require further study. Firstly, our statistical approach used the latitude and longitude of LTLAs to assume a smoothed Euclidian representation of distance across England. This may not be the most appropriate representation for infectious diseases that have varying means of transmission across space, such as transport networks or household/employment networks, and alternative covariates could be chosen to determine links between neighbouring regions. The impact of this choice of spatial distance may be limited when aggregating to a larger spatial scale such as LTLA, compared to 1km grid squares, for example, and this may be a reason for the spatial heterogeneity in the observed transmission rates in this study. Future work will explore this in more detail combined with extended statistical analysis, for example looking at different time periods.

Secondly, in this study and when using Covasim in general, though we have aimed to use the most recent data from the literature to parameterise the model, some questions remain unanswered. For example, we collated evidence to develop a separate vaccine effectiveness model, detailed in [25]. However, vaccine efficacy against onward transmission of the Delta variant and waning protection from both vaccination and infection are both uncertain from current data. Specifically, the version of Covasim used in this study assumed a single antibody waning function for all individuals and all types of immunity, with individual- and immune-level variation in the level of NAbs. As future results reveal differences in the antibody kinetics of natural- vs. vaccine-derived neutralizing antibodies, this assumption will be revisited. Additionally, Covasim is being adapted to capture the effect of heterologous immune sources, including different vaccines administered at different intervals. This topic will continue to grow in relevance as countries roll out vaccine boosters to its populations, and we plan to revisit it in future work.

Thirdly, we calibrated the model to aggregated national data on reported cases, hospitalisations and deaths. In future studies we will look at fitting more granular distribution of these epidemic metrics across different ages, especially when modelling the impact of different vaccination strategies including immunisation of under 18 years old and booster vaccination for elderly people.

Finally, we chose the dates to compare different variants to reflect the period of exponential growth of the emerging variant as per Figure 1, and we only considered LTLAs where the prevalence of the variant reached greater than 50%. This method of temporal truncation is different to the method of [6] in which the transmissibility of Alpha and B.1.177 were compared only over the first 31 days of the emergence of the new variant. In future work we will explore the differences in transmissibility when different periods of studies are used, and different prevalence threshold values for transmission.

In summary, we used statistical analysis and agent-based modelling to quantify the progressive transmissibility of three SARS-CoV-2 variants that have been circulating in England between September 2020 and July 2021, and used this information to model the epidemic in England over this period. Our findings confirm that these new variants became progressively more infectious over time. The emergence of highly infectious Delta variant in late spring 2021 resulted in a one-month delay of Step 4 of England’s COVID-19 roadmap out of lockdown. In our simulations, conducted in June 2021, we found the delay could not prevent but could drastically dampen the projected resurgence due to Delta, which was further suppressed as a result of the age-prioretised vaccination programme from December 2020.

## Data Availability

Publicly available data were used in this study and are available upon reasonable request to the authors.

https://github.com/Jasminapg/Covid-19-Analysis

## 6. Contribution

JPG conceived the study. JPG and BS developed the statistical analysis methodology and BS ran the regression modelling with input from JPG. JPG developed the specific Covasim modelling framework for this study, based on the Covasim model developed by CCK, RMS, KR, JC and DJK. JPG, KR, JC, CCK, RMS and DJK collated data for the parameters used. JPG ran the mathematical modelling analysis with input from KR, RMS and CCK. JPG, RH, LF, CF, RV and CB defined the different scenarios in the UK context following conversations within UK Health Security Agency, Scientific Pandemic Influenza Modelling Group (SPI-M) and Scientific Pandemic Influenza Behaviour Group (SPI-M) and Scientific Advisory Group for Epidemics (SAGE) which give expert advice to the UK Department of Health and Social Care and wider UK Government. JPG wrote the manuscript with input from BS, CW, RH, RMS, JC, FdL, WW, RV, CB, KR, AI, DJK, CF and CCK. All authors approved the final version. JPG is the manuscript’s guarantor.

## 7. Acknowledgements

JPG’s work was supported by funding from the UK Health Security Agency and the UK Department of Health and Social Care (DHSC). This work was also supported by DHSC funding awarded to CF and Li Ka Shing Foundation grant awarded to CF. COG-UK is supported by funding from the Medical Research Council (MRC) part of UK Research Innovation (UKRI), the National Institute of Health Research (NIHR) [grant code: MC_PC_19027], and Genome Research Limited, operating as the Wellcome Sanger Institute. The funders had no role in the study design, data analysis, data interpretation, or writing of the report. The views expressed in this article are those of the authors and not necessarily those of the UK Health Security Agency or the UK Department of Health and Social Care. BS is a member of the Scottish COVID-19 Response Consortium, which was undertaken in part as a contribution to the Rapid Assistance in Modelling the Pandemic (RAMP) initiative, coordinated by the Royal Society.

